# Nothing without connection” – participant perspectives and experiences of mentorship in capacity building in Timor-Leste

**DOI:** 10.1101/2023.06.08.23291064

**Authors:** Jennifer Yan, Nelson Martins, Salvador Amaral, Joshua R Francis, Barbara Kameniar, Clare Delany

## Abstract

**Background:** The literature on mentorship approaches to capacity building in global health is limited. Likewise, there are few qualitative studies that describe mentorship in capacity building in global health from the perspective of the mentors and mentees.

**Methods:** This qualitative study examined the perspectives and experiences of participants involved in a program of health capacity building in Timor-Leste that was based on a side-by-side, in-country mentorship approach. Semi-structured interviews were conducted with 23 participants (including Timorese and Australian mentors, and local Timorese counterparts) from across a range of professional health disciplines, followed by a series of member checking workshops. Findings were reviewed using inductive thematic analysis. Participants were included in review and refinement of themes.

**Results:** Four major themes were identified: the importance of trust and connection within the mentoring relationship itself; the side-by-side nature of the relationship (akompaña); mentoring in the context of external environmental challenges; and the need for the mentoring relationship to be dynamic and evolving, and aligned to a shared vision and shared goals.

**Discussion:** The importance of accompaniment (akompaña) as a key element of the mentoring relationship requires further exploration and study. Many activities in global health capacity building remain focused on provision of training, supervision, and supportive supervision of competent task performance. Viewed through a decolonising lens, there is an imperative for global health actors to align with local priorities and goals, and work alongside individuals supporting them in their vision to become independent leaders of their professions. We propose that placing mentoring relationships at the centre of human resource capacity building programs encourages deep and transformative learning, and is more likely to lead to long term, meaningful and sustainable change.

## Introduction

Despite progress worldwide to improve provision of health care, reduce infant and child mortality, and improve life expectancy and living standards, significant disparities continue to exist in the quality of health services available, and resultant health outcomes between different countries. Global health capacity building work often involves the provision of financial support and technical assistance from well-resourced settings to less well-resourced settings, through initiatives designed to build skills and capacity of the health workforce in order to strengthen the quality of health systems and improve health care delivery.

Strategies utilised for human resource capacity building in global health are many and varied, with examples of overseas training, intensive in-country support, short course training or workshops, and supervision and mentoring for short stints that may be one-off or with recurring follow-up of variable frequency (1–13). In-country training and supervision may be conducted in-situ with existing work teams or with selected staff only in a centralised location, or even remotely via email or phone.

The method and mode of capacity building education has impacts on the quality of the learning and subsequent sustainability of that learning for the individual and the broader health service. Different techniques for teaching and delivering content have implications for the promotion of superficial or deep learning, rote learning versus critical thinking, and successful application from theory to practice (14–17). For example, individuals receiving overseas training in more developed settings may experience a sense of great internal transformation and learning, but struggle to troubleshoot and overcome barriers to change as a solo change agent on returning to their own country context and workplace, where they may be faced with very different issues to those encountered during their training (12,13). Short course trainings or workshops may be less likely to lead to sustained changes in performance in the absence of ongoing support and provision of feedback and follow-up (7,16–20). Remote mentoring and supervision offers flexibility, but can be limited by unreliable access to phone and internet connectivity and greater challenges in developing relationships, avoiding miscommunication, and adapting advice and educational materials to the local context, particularly when parties come from different cultural backgrounds (12,21).

The concept of mentorship is one that is well accepted in the educational literature. The mentor is compared to an advisor or guide, with two way communication and mutual respect between mentor and mentee, and where the relationship is integral, not just for the transfer of information, but also as a means to inspire and guide to transformative learning (22,23). Though the concept of mentorship is well recognised in the field of education, the dominant focus in global health capacity building literature is on task performance and related supervision (1,2). Literature describing the use of mentorship is limited (1,2,6,8).

The theoretical underpinnings of the mentorship approach are, however, closely aligned to the concept of building capacity in a respectful, collaborative way, by encouraging two-way learning and providing opportunities for the mentee grow and lead, not just to learn.

Published literature evaluating mentorship approaches in global health has favoured quantitative assessment and there are few qualitative studies of participant experience (3,24), particularly relating to mentorship in the cross-cultural setting (25–28). There is a need for more research into approaches that place international mentors into resource constrained settings and to identify the factors that contribute to, and impede, productive and successful mentoring relationships from the perspectives of both parties involved.

This research project describes participant experiences of a particular model of human resource capacity building involving intensive in-country, side-by-side mentorship with expatriate and local mentors in Timor-Leste.

Timor-Leste is located in South-East Asia, an island nation sharing a land border with Indonesia. Timor-Leste is a relatively young country, having achieved independence 20 years ago following Indonesian occupation and prior colonisation by the Portuguese (29). Timor-Leste lost approximately one-third of its population during almost three decades of conflict, along with its infrastructure, and a generation of leaders. Timor-Leste is considered a resource-constrained setting, and is ranked 140 out of 189 countries and territories on the Human Development Index (30). Much of the development that has occurred since independence, including the rebuilding of a national health service and health workforce, has occurred with involvement of external development partners (31).

The Menzies School of Health Research (Menzies) is involved in delivering a program of capacity building, research and health system strengthening in Timor-Leste, working in partnership with the Government of Timor-Leste Ministry of Health, and Ministry of Agriculture and Fisheries, via projects funded primarily by the Australian and UK governments. Menzies is an Australian medical research institute dedicated to improving Indigenous, global and tropical health. The Menzies program of work in Timor-Leste has focused on strengthening the country’s local health system capacity to recognise and respond to infectious diseases, which remain a major cause of morbidity and mortality (32–34). The work is centred around a program of intensive side-by-side mentorship with mentors (expatriate Australian and Timorese) working alongside local Timorese colleagues across a range of settings – laboratory, clinical and surveillance; and across a number of professional disciplines in human and animal health - involving doctors, nurses, public health staff, laboratory scientists and technicians, and veterinary staff. The support for human resource capacity building has occurred in conjunction with funding to support improvements in infrastructure, equipment and information management systems. Mentors are based in-country, situated in the workplace alongside their local counterparts, to exchange knowledge and skills, to model, guide, support, and help to troubleshoot application of skills into real life on-the-job practice. The mentorship approach used in this program was informed by theoretical concepts of mentorship from educational literature and a growing understanding of the importance of decolonising global health.

## Methods

### Methodological orientation and theoretical framework

We took an interpretive phenomenological approach to this study. In order to centre participants’ lived experiences, learning from, and with participants, we undertook a collaborative and iterative process of member checking – obtaining, checking and analysing, and drawing meaning from the data, together with participants.

### Study setting

This study was conducted in Timor-Leste. It was designed to examine the mentorship approach of a program of work delivered by Menzies School of Health Research in partnership with the government of Timor-Leste.

The study was conducted between October 2020 and November 2022. Due to the COVID-19 global outbreak, the mode of mentorship delivery changed mid-program as a number of the mentors were unable to continue working in-country. These mentors returned to Australia and continued mentoring remotely. Other mentors were able to remain in-country as their work also involved direct support of the national COVID response. Two Timorese mentors returned to Timor-Leste specifically to support their country and the laboratory efforts at this time. We were also interested to understand the impact of this change on mentors and mentees in their experience of the mentoring relationship.

### Participant recruitment and data collection

We recruited participants who had experience of the Menzies program of work either as mentors or local colleagues. The terminology used by the program was of a mentor and their local colleague or counterpart, rather than ‘mentee’. Potential participants were recruited by first displaying information regarding the study with flyers in the workplace, and mention of the research project at staff meetings, followed by an email distribution to potential participants and a message on a group messaging platform. We planned to undertake purposive sampling, to achieve a mix of participants across professional disciplinary areas and across mentors and mentees. Our initial aim was for 15-20 participants, but we had capacity to include all 23 individuals who expressed interest to participate (8 mentors and 15 counterparts).

We first conducted a series of individual semi-structured interviews, in which we focused on participants’ experiences and perspectives. We then held three follow-up group workshops with participants as part of a member checking process (described under data analysis). The majority of interviews were conducted in-person in a private mutually convenient location, with a few interviews conducted over online videocall. Interviews were planned to be 30-45 minutes in duration, however a number of interviews extended out to 60-90 minutes with participants keen to share their thoughts. Participants were offered the option of being interviewed in English, Tetun, or a combination of both. All interviews were audio-recorded. Interviews in English were conducted by researcher JY. Interviews in Tetun were conducted by researchers NM and SA (fluent in both Tetun and English), and later transcribed verbatim and then translated from Tetun to English, with review of the accuracy of the translation by the research team, and opportunity provided for review, clarification and edits by the participant.

The research team met together on multiple occasions to review the audio recordings and transcripts and reflect on interviews as they were conducted, discussing approaches to encourage natural conversation and ensure participants felt comfortable to share their experiences honestly, including to provide negative feedback. This was predominantly achieved by revisiting the aims and intent of the research project at the beginning of each interview, sharing the motivation for conducting the research, and our desire to learn from and be guided by participant’s experiences and suggestions for how things could be done better, in designing future mentorship and capacity building work.

The data collected included the audio recordings, transcripts and translations of the interviews, and also butcher’s paper notes, video excerpts and photos from the ‘member checking’ group participant meetings described below, recordings and translations of discussions, and notes, observations and reflections from discussions of the research team.

### Data analysis

Transcripts were analysed and coded by the principal investigator JY and reviewed together with discussion of themes by JY, JF, NM and SA, with further discussion and development and refinement of themes with CD and BK. Participant quotes were coded numerically e.g. P3 for participant 3. Because of the small sample size and specificity of roles, a simple numbering system was used that does not distinguish nationality or role, for purposes of preserving anonymity.

Inductive thematic analysis was supported and supplemented by a process of member checking to ensure trustworthiness of the findings (35). This process of member checking involved three half-day meetings where research participants were invited to workshops to reflect upon and discuss key questions raised in the mentorship interviews. Participants were informed that these workshops were being conducted to provide a member checking process for the research data and analysis. The workshops also provided an opportunity for participants to share their experiences, learn more about mentorship, and help steer the organisation’s direction in its approach to mentorship and ongoing capacity building initiatives. Participants consented to the data generated in these member checking workshops being included in the research. Through the member checking process, participants also contributed to the process of thematic analysis, reviewing and refining themes (35).

The methodology chosen was consciously collaborative and participatory, to identify the themes of greatest importance to the participant group as well as differing viewpoints, to limit the impact of views and assumptions imposed by the researchers, and to provide participants a shared forum and pathway for their suggestions to be heard and influence the program’s direction and implementation.

### Ethics

Ethics approval was obtained from the Human Research Ethics Committee of the Northern Territory Department of Health and Menzies School of Health Research (HREC2020-3891) and Instituto Nacional de Saúde in Timor-Leste (1684MS-INS/GDE/X/2020). A participant information sheet was provided in Tetun and English and written informed consent obtained from all participants.

## Results

There were 23 interviews conducted. These included 9 female and 14 male participants, and participants across human (n=18) and animal (n=5) health, from a range of professional disciplines including medical, nursing and pharmacy staff, laboratory scientists, surveillance and public health staff, veterinarians and veterinary technicians. There were six Australian and two Timorese mentors. A number of Timorese participants identified subsequently during the interviews with the roles of both mentor and mentee.

Participants’ answers were frank. They shared both positive and negative experiences and acknowledged many nuances to the relationship and the mentoring experience. Many participants wanted to continue the interviews over the allocated time, and to continue providing their thoughts and suggestions for improvement.

The most prominent theme from the study was the fundamental importance to participants of trust and genuine connection within the mentoring relationship itself, the foundation and centre from which deep engagement and transformative learning was able to take place. Other key themes included the side-by-side nature of the relationship (*akompaña*); mentoring in the context of external environmental challenges; and the need for the mentoring relationship to be dynamic and evolving, aligned to a shared vision and shared goals.

Mentors and their counterparts reflected on similar issues from different standpoints, and often their comments showed insight into the position of the other. There were no clear differences or trends across the professional disciplines in how mentorship was perceived, though experiences of mentoring relationships varied between individuals; the inclusion of a wide interdisciplinary group showed that the nature of mentorship was not specific to profession.

### Theme 1: Importance of trust, and genuine connection in the mentoring relationship - ‘nothing without connection’

The word connection was used often, and from this connection, participants described potential for deeper engagement, an alignment and agreement to partner along a path of development, and deeper rather than superficial learning.

> “Mentorship stuff is about personal approach… So, having someone working closely with you, it gives you, I don’t know, a positive aura. And also… like a guarantee sort of way. Okay, I think I have someone who can help me to achieve this. I think I have someone who has this background knowledge. And I want to go there. And this is the someone. ‘Come with me.’” (P23)

Participants across all areas spoke at length about the importance of the relationship, recognised by mentors as well as local counterparts, as being necessary for learning, motivation, and productive communication. Participants spoke of the necessity of mentorship being not just a professional relationship but individual connection, and of friendship (Figure 1).

**Fig 1.**
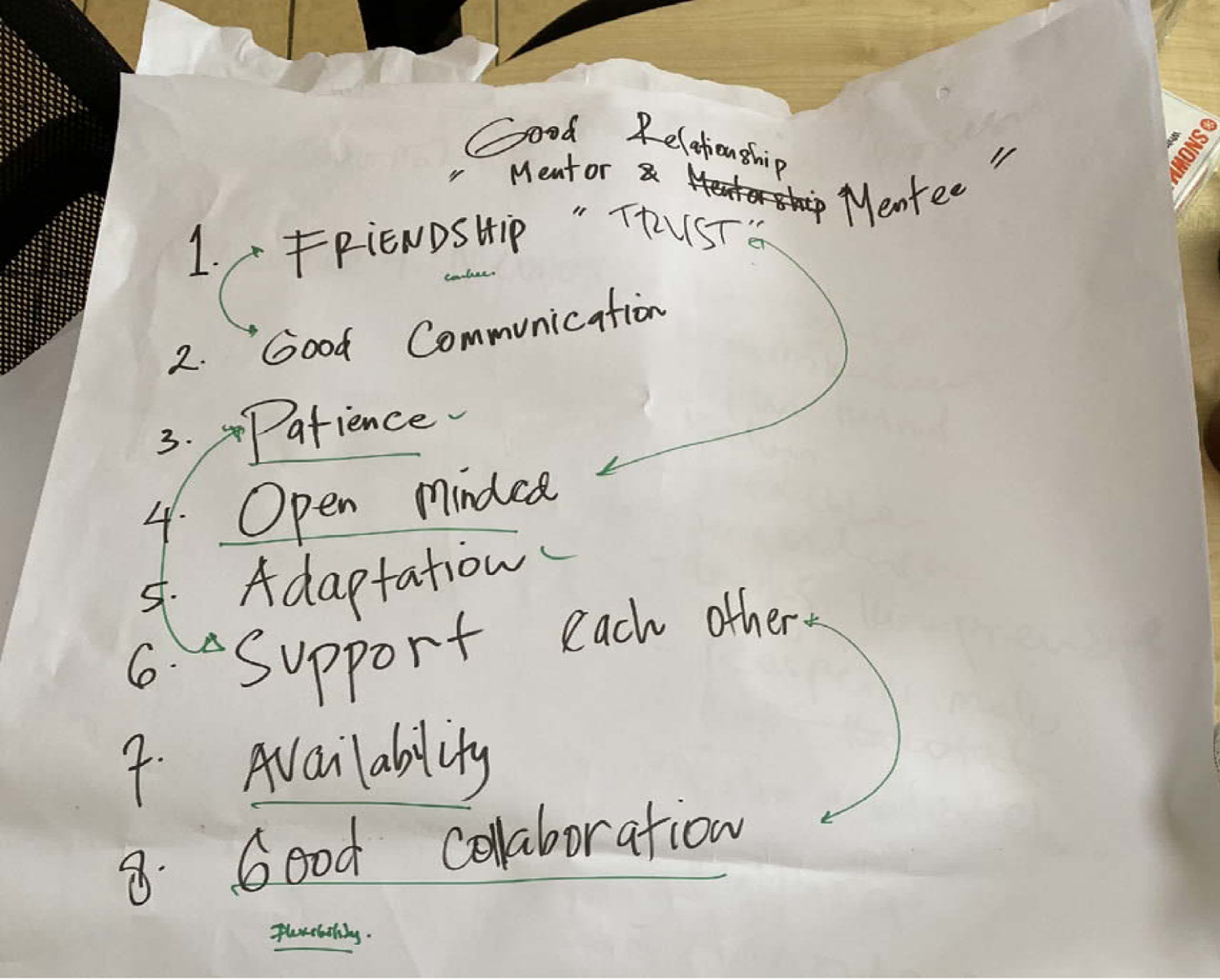
Example butcher’s paper notes from group participant meeting on ‘what makes a good relationship between mentor and mentee’

> “There’s no magic. Just making friends, eating food. The relationships are important… you don’t tick them off, or sign off that the relationships are going well. That’s not a, um, KPI or anything. Building relationships. But it’s important.” (P16)

> “Side-by-side… This type of relationship, there is more confidence… instead of you’re calling doctor, he can be like a brother, when you are too close, you’re able… you are sacrifice yourself to teach someone without return. I mean, without any payment. They respect you.” (P14)

Participants spoke about experiences they had had in training and mentorship programs where this kind of relationship and adaptation to context was not present, and their frustrations, the impact on their learning, motivation, sense of self-worth and achievement.

> “Especially if you come and you are the one who orders, only orders, only to teach, we feel like we are being fooled so that sometimes obstructing… work does not go well.” (P8)

Participants emphasised that the development of this connection takes time. It requires getting to know each other personally.

> “The first five or six months, there wasn’t a lot of output from my behalf, but the first third of the project was just establishing relationships… which is an important part of mentoring. If you take time to do that properly… what’s going on now has been an accumulation of a lot of background work over years and now it’s built to a crescendo where things are firing along”. (P16)

In-person presence, in-place, in-country, working side by side, was how this developed. Mentors reflected on how much better they were able to support their local colleagues once they had this trust and understood the contextual challenges. They were then able to adapt their troubleshooting suggestions appropriately.

The Timorese participants very clearly requested that mentors stay for a year, and ideally longer. In the group meetings, Timorese participants described the burden placed on them when mentors changed frequently, sometimes every six months. Adjusting to the new person and new personality, re-orientating another foreigner to the peculiarities of the local context and cross-cultural differences, introducing them to colleagues, and navigating stakeholder relationships were identified as challenges. Individual mentors would come with slightly different ways of working, and different suggestions, before coming to understand the context and history of what had come before them.

> “These conditions make us feel stressed. Because what we have learnt from first mentor is different, the second come different, the third mentor also different. Sometime it’s difficult. After one mentor came and then another mentor replaced with different system. I understand because even they are all from Australia but they are from different hospital so sometimes it happened.” (P7)

Establishing deep trust required time. Once established though, the impact of this trust on achieving a shared understanding was significant.

> “[x] is quite honest with me. So I get the real story behind things rather than the official story, or the story at face value.” (P16)

The key finding from this study, then, was the importance of trust and connection in the mentoring relationship, and that this trust and connection takes time and needs to take place *in situ*. Participants reflected on the impact of the mentoring relationships on the broader program of work.

> “It’s the glue that held it together that actually made it work. If it’s not for those mentor relationships, we wouldn’t have pulled this off. These machines would have sat in isolation and not been used, and the technology wouldn’t have been incorporated… the understanding of the people that are now going to have to take this on only works because of the mentoring relationship. It would have been nice to foster even more of that to be sure that it will live on.” (P17)

### Theme 2: The side-by-side nature of the relationship - “akompaña”

When asked what mentorship meant, Timorese participants reflected that there was no direct translation for the word mentor or mentorship into Tetun, the local language. Many participants used the word ‘*akompaña’* to define their experience of mentorship, describing a sense of being accompanied, alongside, side-by-side, being motivated and supported by their mentor.

> “He is the one who always motivate us..” (P8)
>
> “Like assisting with the things that we or mentor knows, the one that we do not know and to remind, motivate, give a good way the one who do not know, or to make it deeper to thing that already exist before..”(P11) “More, contact, and to be side by side… I expect to have a mentor who works side by side with me.” (P14)

This was reinforced in the group participant meetings. Timorese participants preferred this word ‘akompaña’ to another word ‘matadalan’ – also a word which means ‘to guide’, but importantly, used in a different sense of being guided ahead and following behind (rather than alongside), as one might have a guide to show you the path up a high mountain. This sentiment was also reflected by the mentors.

> “Well… you’ve got to build a bit of trust don’t you. It’s not like a teacher student relationship, like a superior inferior… is that what you call it? It’s a bit of… sort of side by side. It’s definitely not hierarchical, I’m the boss and they’re the student. Mentoring is side by side. They roll their sleeves up and… like in the lab, (working) on the bench… not just throwing advice from afar.” (P16)

Both Australian and Timorese participants used similar language to describe the nature of the relationship, and it was clear that the concept of mentorship, with elements of mutual respect and two-way learning, translated across cultures.

Australian mentors described needing and receiving *“a kind of cultural mentorship.” (P22)* Participants reflected on the difference between this style of long-term, side-by-side mentorship and other models of teaching and training that they had previous experience of as either teachers or learners. Whilst recognising the role that lectures and workshops could have in disseminating information to a large group of people, both mentors and local counterparts noted the limitations of the group setting on learners feeling able to ask questions, to have teaching pitched to the right level, and on translating knowledge or skills into practice. In the member checking workshops, some Timorese participants proposed that training modalities such as group lectures or workshops would be of greater value if they could be implemented alongside a follow-up schedule of regular, frequent visits for workplace-based training and support, and sharing of contact details for less formal and remote support between times. It was felt that this kind of set-up could utilise the benefits of mass information delivery while still then addressing an individual’s specific learning needs – as long as there was a consistent mentor who they could develop a trusting relationship with over that time. These descriptions essentially articulated and added in key elements of support necessary to achieve the kind of accompaniment that they had experienced.

Participants valued the opportunity through mentorship for the local counterpart to actively direct their learning journey, and described a relevance and depth of learning, that was more difficult to achieve through other forms of training where learning is seen as unidirectional, teachers as active and learners as passive recipients of the ‘wisdom’ of the teacher. Such a disposition to teaching and learning reflects the colonial past of many new nations and is not only ineffective as a pedagogical approach, but also demeaning. Instead, mentorship as ‘akompana’ privileges the relationship as equal and different. It requires active listening, understanding, and responding (Table 1).

**Table 1.** Theme 2: The side-by-side nature of the relationship. Quotes illustrating the concept of “akompaña” and participant views on the benefits of this approach compared to other experiences of teaching, training and supervision.

|  |
| --- |
| **From the mentors** |
| *I see myself as not just teaching or showing the right way to do things, but also showing them our approach, showing them the attitudes and showing them the character of embracing and knowledge sharing and not holding back. (P23)* |
| *The mentoring relationship is 'what don't you know? What do you want to know?' There's a lot more questions and it requires the mentee to come into the relationship to fill their knowledge gaps and to think and to start questioning themselves about what they don't understand and what they want to advance on. (P17)* |
| *I think that's a very important aspect of mentoring, that it's bi-directional and getting the questions back and establishing the hunger for knowledge as well. (P17)* |
| *You sit side-by-side with someone and you understand where they're coming from; you understand their background; you understand their limitations in language or in learning and past experiences and you adapt the teaching model to that particular individual and then you have immediate feedback on whether there's an understanding or a learnt concept. It embeds it.. it embeds it properly and you can adapt what you're doing to each person. I think that model... side-by-side mentoring, which is picking up on holes immediately when they're seen and not just a PowerPoint which apparently covers everything that you're ever supposed to know on that particular topic done in an hour: blasted and never checked again, is the difference. To try and understand what the learners need and want. To go at the pace of the learners and to pick up on ad hoc, opportunistic moments where it's obvious that the learner doesn't quite understand and to feed into those at the point where it's going to be most useful for them. (P17)* |
| *The first thing for a mentor is to listen and to watch and to understand: understand the system; understand the person; understand the limitations and understand what's going on culturally, socially, as well, in that setting. So you can formulate whatever training and whatever response is necessary based on those things. (P17)* |
| *We care about the individual and the individual that we're training may or may not grasp the third step in that seventeen step process and so therefore we need to spend a lot more time on that third step. So there's this adaptable, person-centred approach. (P17)* |
| **From their local counterparts** |
| *They will always be available to accompany, not as formal, we can ask them questions. For mentorship we feel very close. If there is a questions we will feel free to ask, and if there is on the job training, will be shows directly. (P9)* |
| *The one I like the most, is when they came, they asked first, and understand in which part we were weak and wrong, and then observed it, and then they follow [up] one by one. (P9)* |
| *So we felt that there always someone available to backing us up. Even though we felt something really and hard and difficult and many things that we do not know but when we feel that they are standing behind, we feel fine, and confident. (P9)* |
| *When he gives feedback, he gives an opinion regarding our work, he wouldn't make that we felt shy or feel embarrassed or something, so I personally never feel like that, even when we did something not right, he will be talking to us in a good way. I like him because he doesn't make us feel inferior. (P3)* |
| *The one that went well was the flexibility. If we feel uncomfortable, we can speak up, if we have an opinion he encourages to speak up. (P1)* |
| *To be useful in mentorship, teaching someone by doing it is very useful. Not, not just theory, and then this is not useful. It's very wonderful working with them. (P14)* |
| *You mentor.. guiding them to achieve an objective... So, you need to accompany them step by step process. You should keep guiding and if they give up you should bring them up, should keep helping them, keep backup until they achieve the goals that we want. Keep accompany them, improving, developing them. (P5)* |
| *There is a relation with mentor. We accompany. With mentor and mentees. It means that mentor giving assistance to mentees to do some works in order to achieve his or her objective. (P2)* |

For participants, being ‘side-by-side’ was integral to the development and growth of the relationship itself. It also provided the context for teaching and learning through direct observation and feedback in the workplace. Participants expressed the strength of this situated learning approach as one of teaching through showing, and learning through doing, gaining apprenticeship not just of knowledge and skills, but of attitude, critical thinking and role modelling; and with the benefit of immediate application and adaptation to context. This kind of relationship required trust, and time to develop, and was considered a deeper and more personal relationship than that between teacher and learner, or with a regular work supervisor.

However, there were times when language barriers impacted effective communication and therefore the ability to effectively work alongside, and build a trusting, two-way relationship between mentors and their local counterparts. One of the Australian mentors was fluent in Tetun; the others lamented that they had not learnt more of the local language. Many of the Timorese participants interviewed had developed their English skills over time, whereas others had limited English, and were therefore impacted to different degrees.

Participants reflected on the impact of different levels of language ability and the unintended effects this could have of limiting the potential of a mentoring relationship, excluding some people in a group from learning, or creating hierarchies of knowledge and access, or perceived favouritism within a group.

> “Colleagues were complaining us because they felt we that having no barrier to the language are the one more close to the mentor, that is why they take some distance because they feel it is unfair to them. So they feel that the mentor just want to teach only the good one, that is why the colleagues did not want to learn because of the language barrier.” (P9) “We have limitation (in language) but doesn’t mean we don’t (want to) know, we try our best to learn. As I said, language is limiting us to ask more.” (P7)

### Mentors were very aware of the issue

> “The people that are understanding a lot of the mentoring and are part of the continuous mentoring relationship are the ones that have the most proficient English. Others unfortunately aren’t as progressed within the mentoring relationship because of the language difficulty barriers —they’re still part of the mentoring process, but the advances that you can make are limited by that. We’ve tried not to ignore them at any step, but they’re not engaged and it’s purely a language thing, the mentoring relationship, because the language isn’t translated across. It forms teams; you don’t want to have this little, secret mentoring group over here where they find out all the information, whereas the other group doesn’t find out the information.” (P17)

Participants recognised nuance in the relationships too, and the influence of individual personalities on the success of mentorship. Attributes of a good mentor were described as being open, approachable, available, flexible, patient and humble. Timorese counterparts saw that a willingness to be open, ask, questions, and accept new knowledge were important characteristics in order to make the most from the mentoring relationship. Participants described the contrast between an effective mentoring relationship, and other workplace relationships where knowledge may be held as power, instead of shared.

Some participants had worked alongside different mentors, and a few reported variable experiences. Those with less positive mentorship experiences described lacking strong connection, or wishing that their mentor had provided more training and support, like they saw others receiving. Contributing factors raised included different personalities and approaches to communication, a different understanding of what the mentoring relationship was to involve, language barriers, and lack of time.

Participants felt that the best chance of ensuring good mentoring relationships would be through careful selection of mentors with personal qualities that would be a ‘good fit’ not just for mentorship but for working cross-culturally, orientation to the role for both parties with description of intended approach and expectations of mentorship, maintaining the individualised nature of support through regular daily informal communication, regular discussions (less frequently) to reassess goals, and informal peer support of new mentors and mentees through regular group catch ups. This (the current approach) was supported and preferred over adoption of a more formal process using written mentorship agreements or mentorship plans, or a ‘one size fits all’ approach.

### Theme 3: Mentoring in the context of external environmental challenges

Many participants mentioned challenges outside of the mentoring relationship; structural limitations that constrained their operating environment and impacted on what mentoring and support of an individual could achieve. Within country, participants expressed having limited power to effect change within a workplace without buy-in and leadership from supervisors and leaders within the government structures, including when shifts in leadership result in changing agendas and the need to establish new relationships.

> “It’s obviously clouded so much in Timor by the political nuances, the different political groups, which I still don’t quite understand and those divisions which most Timorese people utterly stand by creates conflicts behind the scenes that you don’t understand. But trying to communicate with all different stakeholders… it just can’t be done enough. Having communication and creating communication channels with these people (in senior roles) is just so essential to success in anything. And not maintaining that can mean that you find doing things (not meaning the mentoring relationship) later on are difficult and remain hard.” (P17)

### Limitations of government budget also constrain available spending

> “The government has no money. Each year when budget comes and there is available budget then we can move a bit… but after finished then we also stop, so it’s hard. We are working together with partners from [lists institutions], they sometimes come and help.” (P11)

Other environmental factors can also influence success, especially if mentoring and training is not paired with corresponding investment in infrastructure, equipment or logistical support for people to be able to do their jobs well. Projects are limited by the interests and requirements of grant funders, and by short funding timelines, though an approach centred on mentorship could contribute to longer term sustainability beyond project end.

> “It’s hard with the way things are funded, like [project] is a ridiculous amount of money in a short time, it doesn’t allow for a lot of.. like, you can’t lock in mentoring. Longer term funding cycles are probably much better for this sort of [mentorship] thing, they lock in a bit of stability and longevity. I mean, part of the thing is recognizing that [if] the funding only goes for so long, that then when people develop a relationship, you still carry on supporting someone, regardless of whether there’s money or not.” (P16)

Additionally, the global COVID pandemic led to unavoidable changes in mentorship arrangements; restrictions on overseas travel of Australian citizens meant many Australian mentors had to leave Timor-Leste and change from working in-country to supporting remotely. This had a variable impact on their Timorese counterparts, depending on their level of skill and experience, their need for close supervision and hands on input, and the closeness of the pre-existing mentor relationship. Once the relationship was well established, online communication could still be effective, but remote mentorship was not the same as being there in person. For some, who felt ready and were competent to work independently, the distance worked well in clearly transitioning task ownership and responsibility, whilst still having regular frequent contact allowing them to feel supported and able to ask questions when needed. These participants identified advantages and disadvantages: of stepping up and not becoming dependent on the mentor’s presence and contribution to workload and decision making, but also losing the benefits of having their mentor by their side.

> “So at that time when we need something he directly responding, not even five minutes he quickly responding, so he is also doing video call, we also taking picture and sent him to have a look.. (but) when we need something that we do not yet understand, there is no one come to help for hands on to show.” (P1)

For others, it was difficult for distance not to change the interaction with their mentor, the frequency of communication and sense of being able to ask for support.

> “The distance is making it difficult for me to have contact with my mentor. Well I think it is different. We have a doubt, we will not text it, through the email.. making subsequent questions.. so maybe big doubt, big confusion I will ask, but just asking questions… When we are close by, privately, I can ask you silly questions when we are close.. but [mentor] in Australia and me in Timor is difficult. Yeah I am not bothering them with my silly questions. Have to be big questions.” (P14)

The experience of the participant above highlights how the side-by-side nature of the relationship was important both physically and metaphorically. Despite a relationship that had built over time, and occurred in situ, the strength of the established relationship was challenged and in this instance, disrupted by distance. This participant did feel that perhaps daily, even brief video-calls may have helped to feel more closely connected, as was the experience of their colleague above. Two other participants had had a mostly remote mentoring relationship (following an initial period getting to know each other in person) and described their experience from reciprocal viewpoints. They were in contact multiple times per day and video-called daily. The mentor was very available, and this supported a sense of closeness and connectedness that overcame the distance, such that it felt nearly as though the mentor was present. Inconsistent quality and reliability of internet connection in Timor-Leste, and time zone differences, added an extra layer of challenge for participants engaging in remote mentorship to overcome the sense of distance and communicate effectively.

### Theme 4: The mentoring relationship as dynamic and evolving, aligned to a shared vision and shared goals

Participants spoke of the mentoring relationship needing to be dynamic and evolving as their skills and needs changed.

> “If the person who comes to teach is based on the mapping that he will teach from A to C, but then the mentees who want to be taught their knowledge have reached D, does he have the initiative to teach from F onwards, or they will follow based on one that has been listed? So not limiting us when we want to develop, not limiting our development not feel like because he is the mentor and we do what he tells us to do. So, he gave us an opportunity.. we can developing ourselves and create our own idea.” (P1)

Mentors recognised the need to adjust their initial preconceptions and expectations, and to change their approach and goals to fit the needs and context, not just initially, but along the whole journey.

> “Part of that is a model of also adjusting to what you see on the ground.. what was probably of significant benefit… was (to) just sit and soak up.. and then adjust a bit.. I would have envisioned doing x and more doing y.. adjusting more what we were focussing on.. other topics that were not necessarily high on my agenda going in.” (P22)

From initial periods of listening and observation, identifying gaps and areas for improvement and developing shared goals, participants described how the relationship changed as it progressed through stages of building motivation, understanding the ‘why’ and ‘how’, modelling practice and then gradual advancement in skill toward competent and then independent practice, with changes in the level of supervision required. One mentor described the process of mentoring scientists to perform a complicated laboratory technique to a high quality assured standard.

> “You don’t expect people to learn in the first or second day you know, they take time to change the habit, take time to change the practice. Don’t just say ‘I think you’ve been working on this the last five years. I think you know this’. That’s it, we miss in between. We missed the steps. Slowly showing people instead is the key. Like when we did (it). So he sat down, he did it. I sat down, I did it and compare our results. And takes time.. now they stand on their own now, now we can just stand here and they’ll do it.” (P23)

A very strong theme from both mentors and local counterparts, was the importance of a shared vision and shared goals, being aligned in a desire to achieve the same things. Shared vision and shared goals were necessary between mentor and local counterpart, but were also described by both sets of participants with respect to seeing their personal goals align with those of Menzies, as an organisation with which their work was involved. Both mentors and counterparts linked this with their internal motivations and described strong altruistic reasons for pursuing their line of work, for pursuing further learning and engaging with the opportunity to be involved in mentorship.

Many Timorese participants spoke of a deep motivation to serve their country and people, wanting to improve Timor-Leste, and bring dignity to the Timorese people through their work.

> “What’s motivated me is we can see the process how to help people that [are] sick. What is more important is we can help them. We can give the good results so then we can give a good treatment to the patients. Because in 2006 when the crisis (conflict) was stopped, at that time at first it was only to answer the need in our country when they said there were no analysts, in laboratory and also in districts. So why they open the school and we enrolled. (P7)

A common vision, described in participant interviews and clearly articulated in participant group meetings, was that capacity building work should actively avoid setting up a situation of dependence. Participants (both Australian and Timorese) expressed a strong desire for Timorese to become independent in their practice, in terms of competency to perform tasks independently, and in leadership. Participants referred to the possibility of dynamic and evolving mentoring relationships, which can support a transition to independence. One participant described this eloquently as a progression via small steps from standing alongside, to standing behind, to not being needed.

> “I know it’s small, ah.. he spends time in there every day with the team, and look at them now. Like [x] stood on her own. I know that she still needs help, but they’re, they’re doing a lot of work in pandemic response. And look at micro now. I don’t need to be there. And I know that [x] can deliver, [x] can deliver and others, others can deliver too…” (P23)

Participants expressed their appreciation when there was an explicitly shared goal of establishing independent practice. As the mentoring relationships had evolved over time, a number of Timorese participants actively expressed a desire to move toward more distanced support, and to take the lead themselves.

> “I think their presence here… is very helpful. But on the other hand, we don’t have to depend on them forever. It is important because it doesn’t mean that we ourselves are not capable.. because one day we can. Do everything ourselves. There are things that we can do, we can decide for ourselves. For example, as now they have provided training for our infectious diseases doctor, our infectious diseases doctors are like our two friends. So far, we really need a line of people who know a little about microbiology, and the patients, and if these people are well trained, lab technicians can also make decisions in the lab even though they still need mentors from a distance. I think at times, we need [mentors].. we can do everything ourselves even though we still need them to still accompany.. from afar. However, in my opinion, if in the future, if we already have the best capacity … I think we are holding in a distance like the mentorship.. but I think it still needs time.” (P8)

Developing this shared vision represented respect, interest and investment in the individual. Those who were able to link their internal motivation with their goals from the mentoring relationship found this very motivating and powerful (Figure 2, interview excerpt).

**Fig 2.**
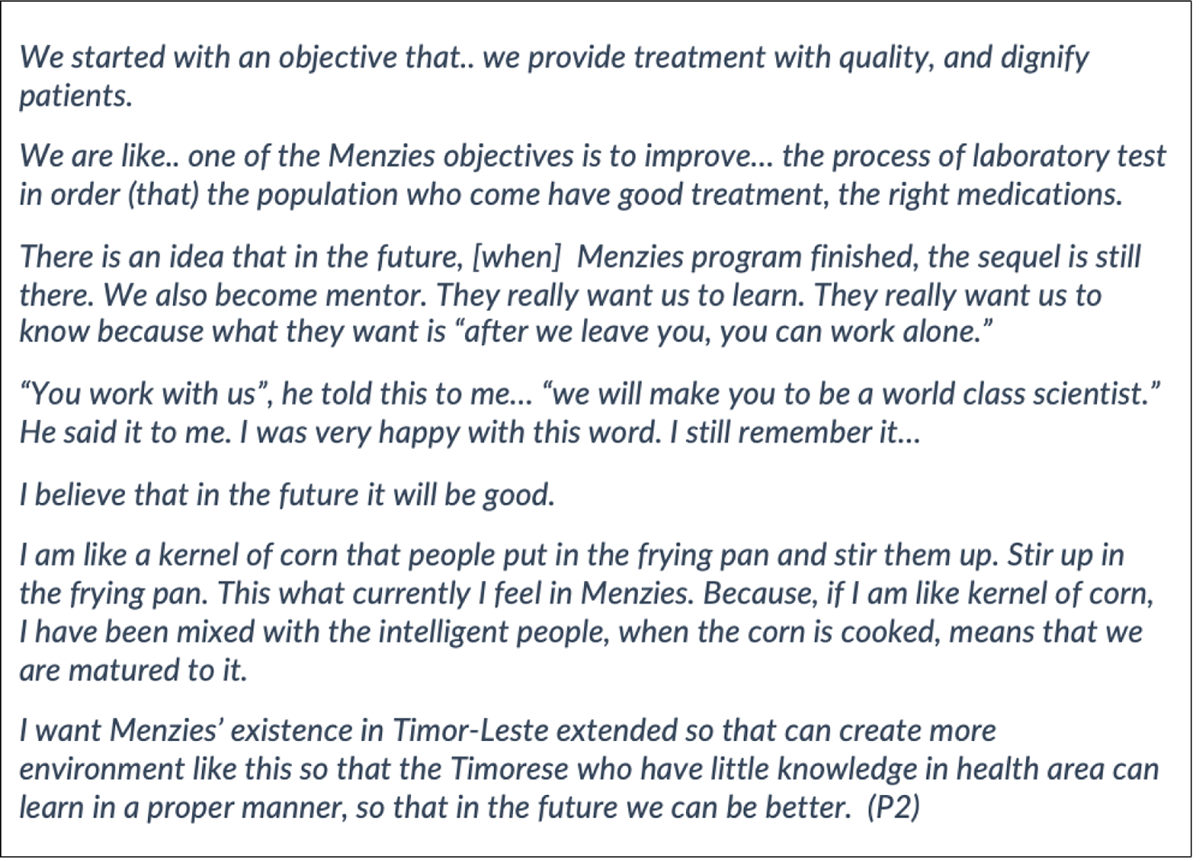
Interview excerpt from a Timorese participant on their personal motivations and a shared vision aligned with their mentor and the organisation.

Participants were frank about the impact of individuals and organisations where respect for Timorese leadership, capacity and vision of a pathway toward independent practice was not demonstrated. One participant described a previous experience in a different program in these terms.

> “And even some people even talk, and I overheard, and even in front of me, like Timor is the easy way to make money because you can just write and copy, paste, and that’s it. Nobody’s checking your report. Because people exploiting the weakness of our institution, none writing technical reports, which the directors won’t even read. Now, I don’t want that to happen to my people.” (P23)

Another shared goal was for Timorese participants to move from being mentee to becoming a mentor themselves. There was recognition of the long-term, sustained change the process enabled, and the potential ripple effect this could have upon others. A number of Timorese participants already identified with both a mentee and mentor role.

> “I define mentees as a learning process to be a good mentor. “What I learned from what he mentored me and what I learned from him, that I taught to other colleagues in the laboratory.” (P9)

Thus dependence and independence were spoken about in terms of task performance but also with reference to Timorese people assuming leadership, taking charge of decision making, and mentoring others. It was important that this was an explicitly shared vision between mentor, local counterpart, and the organisation, and was of particular importance given the country’s history of colonisation and struggle for independence. Some saw the continued influence of external partners, dominating decision-making by the government as a continuation of external control.

## Discussion

The key finding from this study was the importance to participants of connection and trust in the mentoring relationship itself. Side-by-side accompaniment was crucial to development of the relationship, to obtaining a detailed understanding of specific needs and contextual factors, and to be able to provide direct and immediate support and iterative learning in a way that was respectful and founded in genuine care. Participants saw value in a personalised, tailored approach to mentorship, responsive to the needs and strengths of each individual. Our research highlights that a good mentoring relationship was one that was individualised, dynamic and evolved with participants’ changing needs over time. Participants recognised the influence of structural limitations in the broader environment in which mentorship was conducted on what could be achieved. Cross-cultural mentoring relationships benefited from a shared vision and goal of independent practice, of particular significance given Timor-Leste’s history of colonisation and struggle for sovereign independence. The cross-cultural nature of the mentoring relationship and how this was managed was a critical thread, relevant across all themes.

The mentoring relationship provided a basis from which deep engagement and deep learning could take place. Observations regarding the key components of an effective mentoring relationship were consistent with previous descriptions in the literature - open communication and accessibility; goals and challenges; passion and inspiration; caring personal relationship; mutual respect and trust; exchange of knowledge; independence and collaboration; and role modelling (36,37).

We had designed our program of capacity building and health system strengthening work around mentorship as a core component of the theory of change, relying on the transfer of skills and knowledge from mentor to their local counterparts. However, the value of connection in the relationship itself was more significant to participants than we had anticipated. This was highlighted by the quote “*nothing without connection*”. The need for strong trust and connection was even more important in the cross-cultural context; and it was clear that this had to develop over time, and was dependent on a period of in-country presence and the personal characteristics of the mentor.

The potential tensions between mentoring and supervisory roles are well recognised (38,39). Assuming responsibility for program oversight, and monitoring of quality or performance (traditional supervision) can confound the mentoring relationship, and tip the balance from a supportive and encouraging environment to one of monitoring and checking (39). Where possible, having two different people act in the mentor and supervisor roles can relieve this conflict (40). However, in many resource-constrained settings, limitations of human resources can mean that one individual holds dual roles. In Schwerdtle, Morphet and Hall’s model of mentorship, mentorship can be seen as ‘adding’ an additional element of support above and beyond supervision and supportive supervision (2). Our findings add an extra dimension to this understanding, showing that in practice, the dual mentor/supervisor may hold shifting positions from encouraging and working ‘alongside’, to enforcing rules or managing performance as the ‘superior’. In these situations, it becomes all the more vital to maintain a strong connection and open communication to hold this duality of roles well and balance the nature of interactions toward the former rather than latter position.

The style of mentorship participants described as ‘*akompaña’*, learning side-by-side accompanied by one’s mentor, was also illustrative of a number of teaching and learning theories in practice, including situated learning (41), intentional change theory (42), and the ‘zone of proximal development’ in Vygotsky’s sociocultural theory of learning (43). Participants described the benefits of the apprenticeship model of learning, ‘situated’ in the workplace and learning through application and adaptation of theoretical knowledge and skills to real life problems (41). A number of Timorese participants described their personal experience of progress and development through mentorship in words that paraphrase intentional change theory which describes the conscious undertaking to develop a personal vision or goal, identify gaps between one’s current and ideal self, make a plan to move between the two, actively apply oneself to practice, and maintain relationships with people to support you to get there (42). The close ‘*akompaña’* relationship between mentor and their local counterpart, gave mentors opportunity to observe, assess and then regularly adjust their teaching to their counterpart’s needs and level of ability. This resonates with Vygotsky’s concept of teaching to the ‘zone of proximal development’, a space just outside of a learner’s existing capacity and thus the next natural step for advancement, where they are both challenged and supported (43). Danoz’s description of mentorship also uses the intersection between grades of challenge and support to illustrate how optimal growth can occur through mentorship (22). Participants described a process of graded development of competency through different stages of learning, with an inverse correlation to the level of direct/indirect supervision required (44). The end goal of mastery and independent practice was associated in our case with a desire from Timorese participants to shift roles and be able to provide the same mentorship to others.

The model of side-by-side mentorship (accompaniment) as described by our participants and the educational literature is less well represented in the global health literature. Programs of situated, side-by-side mentorship exist in global health but are not the dominant model (2). Anecdotally, a number of non-governmental organisations do engage in models of situated side-by-side mentorship, but their evaluation reports tend to remain confidential internal documents for donors, and articles in the peer-reviewed literature are limited (3,4,13). Discussion in the global health literature on human resource capacity building tends to focus on achieving performance of specific task competencies and thus leans more toward supervision, supportive supervision and training rather than weighting the value of the relationship. Even when the term mentor is used, often interventions themselves are focused on transfer of knowledge and skills without also having a focus on the needs and goals for professional development of the individual (2,45). The concept of ‘*akompaña’* requires of the relationship something that extends beyond the focus areas of a particular project, or even the limited timeframe for which guidance is required. A relationship of accompaniment implies an ongoing closeness and trust, and a connection that might be expected to remain beyond the life of the project.

A number of participants spoke of structural barriers within their environment, outside of the mentoring relationship, related to government, finances and the political landscape, and limitations of the donor funding model; challenges that are recognised globally and also specifically in the Timor-Leste context, in building human resource capacity in health (46–50). In both interviews and group discussions, the importance of investments that address both human resources and training, and infrastructure and equipment, was highlighted, to enable participants to put skills into practice, and avoid a situation where system barriers overshadow any potential gains from mentorship and training (24,51–53).

In our study, the timing of the COVID pandemic facilitated a transition towards independence through relocation of mentors and forced indirect supervision; the varied reactions to this change depended on the strength of the pre-existing relationship and individual levels of competence and readiness for the change. It also provided opportunities for Timorese mentors to assume leadership roles in Timor-Leste, with expanded scope of influence and opportunities to mentor multiple individuals. They were an example of local leadership and were inspirational mentors to many; their input was essential to the national health system’s public health and laboratory response to COVID-19, and their leadership exemplified the theme of a shared vision for Timorese independence that was identified in this research (34).

We found that participants’ understanding of good mentorship aligned particularly strongly with the concepts of transformational teaching and transformational leadership. Transformational teaching and leadership promotes an approach to learning that is linked with deep internal drivers, which can be more powerful than external incentives in motivating change and stimulating deep (rather than superficial) learning (54). Timorese participants spoke of their internal motivators: their desires to learn and improve in order to better serve their country and people, their desires to excel and progress in their careers, to perform their roles independently, and to lead and become mentors to others. The mentors, both Timorese and Australian, described having these same goals for those they worked alongside. Both parties (mentors and their counterparts) described a process of accompaniment on a path toward a shared vision, and described a strong drive in their work when their internal motivators aligned with the goals of mentorship. This kind of alignment is also discussed in the organisational culture literature, where individuals who feel that their aspirations align with those of the organisation, especially when this happens in a bottom-up approach, are motivated by this in their work. Senge defines shared vision as “the capacity to hold a shared picture of the future we seek to create. When there is a genuine vision, people excel and learn not because they are told to, but because they want to” (55). This type of support stands in contrast to other experiences described by participants, of people in leadership positions holding onto ‘knowledge as power’. And for many, these contrasting experiences have motivated a desire to share this approach to mentorship with others.

### Cross-cultural dynamics

Our findings build on what was already known, to show that the concept of mentorship along with other theories of teaching and learning can translate across cultures (56), and from the educational literature to the global health capacity building context. The descriptions given by participants derived from their lived experience of mentoring relationships rather than learnt definitions of theories of mentorship from the literature. It was reassuring to see that our conceptualisation of mentorship in program design had largely been realised in the implementation, and the principles of effective mentorship had carried through. It was evident however, that some participants had had variable and less positive experiences, sometimes related to external factors and at other times related to factors exerting influence within the mentoring relationship - the ‘fit’ between mentor and local counterpart, personal attributes relevant to adapting to work in a lower resource setting, communication style, and language. There were personal characteristics that were noted as advantageous to providing mentorship in a cross-cultural setting. Patience, flexibility, adaptability, and the ability to observe and listen (rather than jump in with an opinion or a fix) were attributes emphasised more by our participants than in previous literature (25,26,36,37,57,58).

Though the concepts of mentorship translated across settings, cross-cultural and contextual differences highlight the need for bidirectional learning and in particular, recognition of the cultural mentorship that participants from a receiving country provide. The concept of cultural mentorship is one that has recently gained more prominence, used in Australia particularly in reference to engaging with Aboriginal and Torres Strait Islander peoples, and respecting Indigenous knowledge (59). Cultural mentorship in the health care setting has been used to build greater understanding of cultural context of the local community, and to support practitioners to provide culturally safe clinical practice, that benefits both the practitioner and the local community (59,60). For Timorese participants in this study, providing cultural mentorship allowed them to optimise the relationship and their own learning opportunities, by giving mentors an appreciation of context, and awareness of how and why they would need to adapt their usual practice, behaviour and communication; mentors in turn were challenged and extended by needing to apply their expertise in new ways and to reconsider their approach.

### A decolonising lens

Understanding participants’ experiences through a decolonising lens (61–63) gives additional perspective to participants’ reflections that for Timorese people, ‘we don’t like to be ordered about or told what to do’, and of the resistance of inaction when subjugated or disrespected, saying ‘yes, yes’ but choosing not to follow through with a plan that does not include shared aims. It was noted that there was a hierarchy sometimes implicit, sometimes overt, of deferring to a non-Timorese. Recalling the quote of one participant who bemoaned money spent on fly-in fly-out international technical advisors and copy-paste technical reports, participants were dismayed to see practices they perceived as blatantly taking advantage of their country, and of the ongoing injustice of people telling others what they should be doing in their own country.

The decolonisation of global health has gained traction in the global health space over recent years, “a movement that fights against ingrained systems of dominance and power in the work to improve the health of populations, whether this occurs between countries, including between previously colonising and plundered nations, and within countries” (64). The concept of global health emerged in response to disproportional health needs between low and high income countries due to resource inequity. However, this positively framed narrative belies the reality that this disparity often exists because of a history of colonisation and exploitation of peoples, states and their resources (65), and an increasing realisation that “the operations of many organisations active in global health… perpetuate the very power imbalances they claim to rectify, through colonial and extractive attitudes, and policies and practices that concentrate resources, expertise, data and branding within high-income country (HIC) institutions” (64).

In undertaking this study, and associated work in Timor-Leste, we are mindful of the importance of decolonisation in our own setting, in our individual interactions and also in the systems and processes that institutionalise power imbalances, that we have power to influence and rectify. The findings of this study give us opportunity to reflect, and further commit to a deliberate, decolonising approach to our work in Timor-Leste. As Timorese and non-Timorese leaders within our organisation, it is essential that we listen, creating spaces where honest conversations can happen, that we recognise privilege, and acknowledge where things may not have been done the best way, accepting the discomfort this brings, while we work to shift power balances (64). As we look to continue creating and maintaining mentoring relationships as part of our work, mentors should work alongside local counterparts, supporting progression to independence and leadership, rather than position themselves for exertion of authority. This adds an extra layer of complexity in instances where mentors also hold supervisory roles. Hopefully, the mentorship of local Timorese into positions of leadership, alongside ongoing respectful partnerships for capacity building and health system strengthening, can contribute to this goal (66).

### Limitations

There were limitations to our study. The dual roles of researchers on this study also working within the Menzies program of work could have contributed to a sense of coercion to participate, positive bias in participant responses, and/or positively biased selection or interpretation of data in favour of reporting good participant experiences of mentorship. However, participants were frank in their responses and gave both positive and negative feedback; a number of interviews ran overtime due to participants wanting to continue adding more reflections, and the group discussions were lively. Participants were a self-selected group who volunteered to participate. Many had close interactions with the program and through their mentoring relationships, were known to the researchers and may have been more likely to provide positive responses. We made efforts to counter the potential for unconscious bias in thematic analysis with a participatory process, member checking, involving participants in revising and refining themes and reinforcing concepts of importance to the participants. Some interviews were conducted remotely via videocall rather than in-person. The COVID pandemic interrupted progress on the research study and so some interviews were conducted at different time points through the transition between in-person and periods of remote mentorship, that could have influenced participant’s feelings about their experience.

## Conclusions

This study builds on limited published literature regarding mentorship approaches to capacity building in global health settings. Capacity building programs that focus on knowledge and skills transfer without broader considerations of teaching and learning theory and the individual needs of participants, can lead to interventions that are efficient to deliver but may result in superficial learning, poor engagement, slow transition to independent practice, and limited long-term sustainable change. Global health human resource capacity building programs should consider incorporation of the concept of accompaniment (akompaña) into program design, with a focus on development of trusting, long-term, side-by-side mentoring relationships, which prioritise development of a shared vision of professional goals.

## Data Availability

Primary data are available from the corresponding author on request.

## References

1. Vasan A, Mabey DC, Chaudhri S, Epstein HAB, Lawn SD. Support and performance improvement for primary health care workers in low- and middleincome countries: A scoping review of intervention design and methods. Vol. 32, Health Policy and Planning. Oxford University Press; 2017. p. 437–52.

2. Schwerdtle P, Morphet J, Hall H. A scoping review of mentorship of health personnel to improve the quality of health care in low and middle-income countries. Global Health. 2017 Oct 3;13(1):77.

3. Manzi A, Magge H, Hedt-Gauthier BL, Michaelis AP, Cyamatare FR, Nyirazinyoye L, et al. Clinical mentorship to improve pediatric quality of care at the health centers in rural Rwanda: a qualitative study of perceptions and acceptability of health care workers. BMC Health Serv Res [Internet]. 2014 Jun 20 [cited 2020 Jun 30];14(1):275. Available from: http://www.ncbi.nlm.nih.gov/pubmed/24950878

4. Catton HN. Developing a mentorship program in Laos. Front Public Heal. 2017;5(6):145.

5. Decorby-Watson K, Mensah G, Bergeron K, Abdi S, Rempel B, Manson H. Effectiveness of capacity building interventions relevant to public health practice: A systematic review. Vol. 18, BMC Public Health. BioMed Central Ltd.; 2018. p. 684.

6. Manzi A, Hirschhorn LR, Sherr K, Chirwa C, Baynes C, Awoonor-Williams JK, et al. Mentorship and coaching to support strengthening healthcare systems: Lessons learned across the five Population Health Implementation and Training partnership projects in sub-Saharan Africa. BMC Health Serv Res. 2017 Dec 21;17.

7. Magge H, Anatole M, Cyamatare FR, Mezzacappa C, Nkikabahizi F, Niyonzima S, et al. Mentoring and quality improvement strengthen integrated management of childhood illness implementation in rural Rwanda. Arch Dis Child [Internet]. 2015 Jun 1 [cited 2020 Jun 30];100(6):565–70. Available from: http://www.ncbi.nlm.nih.gov/pubmed/24819369

8. Feyissa GT, Balabanova D, Woldie M. How Effective are Mentoring Programs for Improving Health Worker Competence and Institutional Performance in Africa? A Systematic Review of Quantitative Evidence. J Multidiscip Healthc [Internet]. 2019 [cited 2020 Jun 30];12:989–1005. Available from: http://www.ncbi.nlm.nih.gov/pubmed/31824166

9. Reddy P, Desai R, Sifunda S, Chalkidou K, Hongoro C, Macharia W, et al. “You Travel Faster Alone, but Further Together”: Learning From a Cross Country Research Collaboration From a British Council Newton Fund Grant. Int J Heal policy Manag. 2018 Nov;7(11):977–81.

10. Kennedy HP, Stalls S, Kaplan LK, Grenier L, Fujioka A. Thirty years of global outreach by the American College of Nurse-Midwives. MCN Am J Matern Child Nurs. 2012 Sep;37(5):290–7.

11. Matovu J, Wanyenze R, Mawemuko S, Okui O, Bazeyo W, Serwadda D. Strengthening health workforce capacity through work-based training. BMC Int Health Hum Rights. 2013;13(8).

12. Guest GD, Scott DF, Xavier JP, Martins N, Vreede E, Chennal A, et al. Surgical capacity building in Timor-Leste: a review of the first 15 years of the Royal Australasian College of Surgeons-led Australian Aid programme. ANZ J Surg. 2017 Jun 1;87(6):436–40.

13. Watters DA, McCaig E, Nagra S, Kevau I. Surgical training programmes in the South pacific, Papua new Guinea and timor leste. Vol. 106, British Journal of Surgery. John Wiley and Sons Ltd; 2019. p. e53–61.

14. Forsetlund L, Bjørndal A, Rashidian A, Jamtvedt G, O’Brien MA, Wolf F, et al. Continuing education meetings and workshops: effects on professional practice and health care outcomes. Cochrane database Syst Rev [Internet]. 2009 [cited 2023 Mar 4];2009(2). Available from: https://pubmed.ncbi.nlm.nih.gov/19370580/

15. Knowles MS (Malcolm S, Holton EF, Swanson RA. The adult learnerL: the definitive classic in adult education and human resource development [Internet]. Elsevier; 2011 [cited 2018 May 13]. 406 p. Available from: https://eds-a-ebscohost-com.ezp.lib.unimelb.edu.au/eds/detail/detail?vid=13&sid=3ce17dfb-7912-4ea6-8441-1529df601e18%40sessionmgr4009&bdata=JnNpdGU9ZWRzLWxpdmUmc2NvcGU9c2l0ZQ%3D%3D#AN=edshlc.013360334-2&db=edshlc

16. Kok MC, Dieleman M, Taegtmeyer M, Broerse JEW, Kane SS, Ormel H, et al. Which intervention design factors influence performance of community health workers in low- and middle-income countries? A systematic review. Health Policy Plan. 2015 Nov 1;30(9):1207–27.

17. Hill Z, Dumbaugh M, Benton L, Källander K, Strachan D, ten Asbroek A, et al. Supervising community health workers in low-income countries--a review of impact and implementation issues. Glob Health Action. 2014;7:24085.

18. Pariyo GW, Gouws E, Bryce J, Burnham G. Improving facility-based care for sick children in Uganda: training is not enough. Health Policy Plan [Internet]. 2005 [cited 2023 Mar 4];20 Suppl 1(SUPPL. 1). Available from: https://pubmed.ncbi.nlm.nih.gov/16306071/

19. Workneh G, Scherzer L, Kirk B, Draper HR, Anabwani G, Wanless RS, et al. Evaluation of the effectiveness of an outreach clinical mentoring programme in support of paediatric HIV care scale-up in Botswana. AIDS Care - Psychol Socio-Medical Asp AIDS/HIV [Internet]. 2013 Jan 1 [cited 2020 Jun 30];25(1):11–9. Available from: https://pubmed.ncbi.nlm.nih.gov/22533352/

20. Rowe AK, Rowe SY, Peters DH, Holloway KA, Chalker J, Ross-Degnan D. Effectiveness of strategies to improve health-care provider practices in low-income and middle-income countries: a systematic review. Lancet Glob Heal. 2018 Nov;6(11):e1163–75.

21. Short J, McDonald S, Turner T, Martis R, SEA-ORCHID Study Group. Improving capacity for evidence-based practice in South East Asia: evaluating the role of research fellowships in the SEA-ORCHID Project. BMC Med Educ. 2010 May 22;10(1):37.

22. Daloz L. Effective teaching and mentoring: Realizing the Transformational Power of Adult Learning Experiences. Jossey-Bass; 1986. 256 p.

23. Mezirow J. Fostering Critical Reflection in Adulthood: A Guide to Transformative and Emancipatory Learning. San Francisco: Jossey-Bass; 1990.

24. Morgan MC, Dyer J, Abril A, Christmas A, Mahapatra T, Das A, et al. Barriers and facilitators to the provision of optimal obstetric and neonatal emergency care and to the implementation of simulation-enhanced mentorship in primary care facilities in Bihar, India: a qualitative study. BMC Pregnancy Childbirth. 2018 Oct;18(1):420.

25. Whitehurst JL, Rowlands J. Helping palliative care healthcare professionals get the most out of mentoring in a low-income country: a qualitative study. BMC Palliat Care [Internet]. 2016 Nov 4 [cited 2020 Jun 28];15(1):90. Available from: https://pubmed-ncbi-nlm-nih-gov.www.ezpdhcs.nt.gov.au/27809819/

26. Parekh N, Sawatsky AP, Mbata I, Muula AS, Bui T. Malawian impressions of expatriate physicians: A qualitative study. Malawi Med J. 2016 Jun;28(2):43–7.

27. Beckett A, Fowler R, Adhikari N, Hawryluck L, Razek T, Tien H. Medical mentorship in Afghanistan: How are military mentors perceived by Afghan health care providers? Can J Surg [Internet]. 2015 Jun 1 [cited 2020 Jun 30];58(3):S98–103. Available from: /pmc/articles/PMC4467500/?report=abstract

28. Finley GA, Forgeron P, Arnaout M. Action Research: Developing a Pediatric Cancer Pain Program in Jordan. J Pain Symptom Manage [Internet]. 2008 Apr 1 [cited 2020 Jun 30];35(4):447–54. Available from: http://www.jpsmjournal.com/article/S0885392407007361/fulltext

29. Government of Timor-Leste. Timor-Leste History [Internet]. [cited 2020 Jul 1]. Available from: http://timor-leste.gov.tl/?p=29&lang=en

30. United Nations Development Programme. Human Development Report 2021/2022. Uncertain Times, Unsettled Lives: Shaping our Future in a Transforming World [Internet]. New York; 2022 [cited 2022 Nov 5]. Available from: https://hdr.undp.org/system/files/documents/global-report-document/hdr2021-22pdf_1.pdf

31. Mercer MA, Thompson SM, De Araujo RM. The role of international NGOs in health systems strengthening: The case of timor-leste. Int J Heal Serv. 2014 Jan 1;44(2):323–35.

32. Sarmento N, Oakley T, da Silva ES, Tilman A, Monteiro M, Alves L, et al. Strong relationships between the Northern Territory of Australia and Timor-Leste. Microbiol Aust [Internet]. 2022 Sep 27 [cited 2023 Jan 14];43(3):125–9. Available from: https://www.publish.csiro.au/ma/MA22039

33. Francis JR, Sarmento N, Draper ADK, Marr I, Ting S, Fancourt N, et al. Antimicrobial resistance and antibiotic use in Timor-Leste: building surveillance capacity with a One Health approach. Commun Dis Intell [Internet]. 2020 Jan 15 [cited 2023 Jan 14];44. Available from: https://pubmed.ncbi.nlm.nih.gov/31940450/

34. Sarmento N, Soares da Silva E, Barreto I, Ximenes JC, Angelina JM, Correia DM, et al. The COVID-19 laboratory response in Timor-Leste; a story of collaboration. Lancet Reg Heal Southeast Asia [Internet]. 2023 Apr [cited 2023 Feb 20];11:100150. Available from: https://pubmed.ncbi.nlm.nih.gov/36744276/

35. Lincoln YS, Guba EG. What is Trustworthiness? In: Naturalistic Inquiry. SAGE Publications; 1985. p. 291–4.

36. Eller LS, Lev EL, Feurer A. Key components of an effective mentoring relationship: a qualitative study. 2013;

37. Hamer DH, Hansoti B, Prabhakaran D, Huffman MD, Nxumalo N, Fox MP, et al. Global health research mentoring competencies for individuals and institutions in low-and middle-income countries. Am J Trop Med Hyg. 2019;100:15–9.

38. Ssemata AS, Gladding S, John CC, Kiguli S. Developing mentorship in a resource-limited context: a qualitative research study of the experiences and perceptions of the makerere university student and faculty mentorship programme. BMC Med Educ. 2017 Jul 14;17(1):123.

39. World Health Organisation. WHO | WHO recommendations for clinical mentoring to support scale-up of HIV care, antiretroviral therapy and prevention in resource-constrained settings [Internet]. 2006 [cited 2020 Jun 30]. Available from: https://www.who.int/hiv/pub/meetingreports/clinicalmentoring/en/

40. Mellon A, Murdoch-Eaton D. Supervisor or mentor: Is there a difference? Implications for paediatric practice. Arch Dis Child. 2015 Sep 1;100(9):873–8.

41. Billett S. Situating Learning in the Workplace: Having Another Look at Apprenticeships. Ind Commer Train [Internet]. 1994 [cited 2018 May 13];26(11):9–16. Available from: http://hdl.handle.net/10072/11865

42. Boyatzis RE. An overview of intentional change from a complexity perspective. J Manag Dev. 2006;25(7):607–23.

43. VygotskiilJ LS (Lev S. Mind in societyL: the development of higher psychological processes [Internet]. Cambridge: Harvard University Press; 1978 [cited 2019 May 3]. 159 p. Available from: http://www.hup.harvard.edu/catalog.php?isbn=9780674576292

44. Griffin P. Assessment for teaching. Assessment for Teaching. Cambridge: Cambridge University Press; 2018.

45. Vasan A, Mabey D, Chaudhri S, Brown Epstein H, Lawn S. Support and performance improvement for primary healthcare workers in low- and middle-income countries: a scoping review of intervention design and methods. Health Policy Plan [Internet]. 2017 [cited 2020 Jun 30];32(3). Available from: https://pubmed-ncbi-nlm-nih-gov.www.ezpdhcs.nt.gov.au/27993961/

46. Cabral J, Dussault G, Buchan J, Ferrinho P. Scaling-up the medical workforce in Timor-Leste: Challenges of a great leap forward. Soc Sci Med. 2013 Nov;96:285–9.

47. Ferrinho P, Valdes AC, Cabral J. The experience of medical training and expectations regarding future medical practice of medical students in the Cuban-supported Medical School in Timor-Leste. Hum Resour Health [Internet]. 2015 Mar 28 [cited 2019 Nov 9];13(1):13. Available from: http://www.ncbi.nlm.nih.gov/pubmed/25880331

48. Hou X, Witter S, Zaman RU, Engelhardt K, Hafidz F, Julia F, et al. What do health workers in Timor-Leste want, know and do? Findings from a national health labour market survey. Hum Resour Health [Internet]. 2016 Nov 18 [cited 2020 Jun 16];14(1):69. Available from: http://www.ncbi.nlm.nih.gov/pubmed/27863499

49. Bertone MP, Martins JS, Pereira SM, Martineau T, Alonso-Garbayo A. Understanding HRH recruitment in post-conflict settings: An analysis of central-level policies and processes in Timor-Leste (1999-2018). Hum Resour Health [Internet]. 2018 Nov 29 [cited 2022 Oct 16];16(1):1–12. Available from: https://human-resources-health.biomedcentral.com/articles/10.1186/s12960-018-0325-5

50. Witter S, Falisse JB, Bertone MP, Alonso-Garbayo A, Martins JS, Salehi AS, et al. State-building and human resources for health in fragile and conflict-affected states: Exploring the linkages. Vol. 13, Human Resources for Health. BioMed Central Ltd.; 2015.

51. Sayed S, Cherniak W, Lawler M, Tan SY, El Sadr W, Wolf N, et al. Improving pathology and laboratory medicine in low-income and middle-income countries: roadmap to solutions. Lancet (London, England). 2018 May;391(10133):1939–52.

52. Potter C, Brough R. Systemic capacity building: A hierarchy of needs. Health Policy Plan. 2004;19(5):336–45.

53. Heller DJ, Kumar A, Kishore SP, Horowitz CR, Joshi R, Vedanthan R. Assessment of Barriers and Facilitators to the Delivery of Care for Noncommunicable Diseases by Nonphysician Health Workers in Low- and Middle-Income Countries: A Systematic Review and Qualitative Analysis. JAMA [Internet]. 2019 Dec 2 [cited 2020 Jun 28];2(12):e1916545. Available from: https://pubmed-ncbi-nlm-nih-gov.www.ezpdhcs.nt.gov.au/31790570/

54. Slavich GM, Zimbardo PG. Transformational Teaching: Theoretical Underpinnings, Basic Principles, and Core Methods. Educ Psychol Rev [Internet]. 2012 Dec [cited 2022 Oct 16];24(4):569–608. Available from: https://pubmed.ncbi.nlm.nih.gov/23162369/

55. Senge PM. The fifth discipline: The art and practice of the learning organization. New York: Currency Doubleday; 1990. 191–215 p.

56. Lescano AG, Cohen CR, Raj T, Rispel L, Garcia PJ, Zunt JR, et al. Strengthening mentoring in low-and middle-income countries to advance global health research: An overview. Vol. 100, American Journal of Tropical Medicine and Hygiene. American Society of Tropical Medicine and Hygiene; 2019. p. 3–8.

57. Gandhi M, Raj T, Fernandez R, Rispel L, Nxumalo N, Lescano AG, et al. Mentoring the mentors: Implementation and evaluation of four fogarty-sponsored mentoring training workshops in low-and middle-income countries. Am J Trop Med Hyg [Internet]. 2019 Nov 14 [cited 2020 Feb 18];100(Suppl 1):20–8. Available from: http://www.ajtmh.org/content/journals/10.4269/ajtmh.18-0559

58. Huybrecht S, Loeckx W, Quaeyhaegens Y, De Tobel D, Mistiaen W. Mentoring in nursing education: Perceived characteristics of mentors and the consequences of mentorship. Nurse Educ Today. 2011 Apr 1;31(3):274–8.

59. General Practice Education and Training Limited AGD of H. Cultural Educators and Cultural Mentors: Building trust and respect [Internet]. 2014 [cited 2022 Oct 22]. Available from: https://aboriginalhealth.gpsynergy.com.au/wp-content/uploads/2021/03/CE_CM-Building-trust-and-respect_.pdf

60. Reath J, Abbott P, Kurti L, Morgan R, Martin M, Parry A, et al. Supporting aboriginal and Torres Strait islander cultural educators and cultural mentors in Australian general practice education. BMC Med Educ. 2018 Oct 11;18(1).

61. Thiong’O NW. Decolonising the mind. Diogenes [Internet]. 1998 Jul 26 [cited 2022 Nov 5];46(184):101–4. Available from: https://journals.sagepub.com/doi/abs/10.1177/039219219804618409

62. Fanon F. Black Skin, White Masks [Internet]. Grove Paperback; 2008 [cited 2022 Nov 5]. Available from: https://groveatlantic.com/book/black-skin-white-masks/

63. Gandhi L. Postcolonial theoryL: a critical introduction [Internet]. Second edition. Columbia University Press; 1998 [cited 2022 Nov 5]. 200 p. Available from: http://cup.columbia.edu/book/postcolonial-theory/9780231112734

64. Khan M, Abimbola S, Aloudat T, Capobianco E, Hawkes S, Rahman-Shepherd A. Decolonising global health in 2021: a roadmap to move from rhetoric to reform. BMJ Glob Heal [Internet]. 2021 Mar 1 [cited 2022 Oct 17];6(3):e005604. Available from: https://gh.bmj.com/content/6/3/e005604

65. Kwete X, Tang K, Chen L, Ren R, Chen Q, Wu Z, et al. Decolonizing global health: what should be the target of this movement and where does it lead us? Glob Heal Res Policy [Internet]. 2022 Dec 1 [cited 2022 Oct 17];7(1):1–6. Available from: https://ghrp.biomedcentral.com/articles/10.1186/s41256-022-00237-3

66. Mogaka OF, Stewart J, Bukusi E. Why and for whom are we decolonising global health? Lancet Glob Heal [Internet]. 2021 Oct 1 [cited 2022 Oct 17];9(10):e1359–60. Available from: http://www.thelancet.com/article/S2214109X2100317X/fulltext

